# Prevalence and spectrum of congenital heart disease in individuals with distal chromosome 22q11.22-23 deletions

**DOI:** 10.1101/2025.10.15.25337915

**Authors:** Tanner J. Nelson, Daniel E. McGinn, T. Blaine Crowley, Lydia Rockart, Audrey Green, Victoria Giunta, Oanh Tran, Daniella Miller, Jeroen Breckpot, Ann Swillen, M. Cristina Digilio, Marta Unolt, Carolina Putotto, Federica Pulvirenti, Bruno Marino, Beverly S. Emanuel, Elaine H. Zackai, Zhengdong D Zhang, Elizabeth Goldmuntz, Erik Boot, Anne S. Bassett, Bernice E. Morrow, Donna M. McDonald-McGinn

**Author notes:** **ETHICS APPROVAL STATEMENT** Our research on 22q11.2 disorders is part of an IRB approved program at Albert Einstein College of Medicine (IRB #1999-201), Children’s Hospital of Philadelphia (IRB 07-5352), University Hospitals Leuven BE (IRB S52418), University of Toronto affiliated hospitals (IRB CAMH 114-2001, CAMH 154-2002, CAMH 150-2002, CAMH 151-2002 and UHN 98-E156), Sapienza (Protocol Ref. 5803) and Rome, IT (Prot. 468/12). A waiver for formal approval under the Medical Research Involving Human Subjects Act (WMO) was obtained from the IRB of Amsterdam UMC, the Netherlands (#W20_098). These subjects were ascertained by their informed consent or assent. All individuals in this study were de-identified for this study.

## Abstract

This study is aimed to determine the spectrum of congenital heart disease associated with distal 22q11.22-23 deletions flanked by low copy repeats, LCR22 D-H. We analyzed cardiology findings in 128 unrelated individuals with distal LCR22 D-H deletions. A total of 62 were newly described and 66 were derived from previous reports. We found that deletions which included LCR22-D as the proximal endpoint were the most prevalent in the cohort (104/128, 81.3%). Clinically relevant congenital heart disease was identified in 48 individuals (37.5%, 95% CI 29-46%), which is lower than the prevalence reported for typical, proximal LCR22 A-D deletions (p=3.7E-4), especially for conotruncal defects (13/128, 10.2%; p=7.1E-13). Mild to moderate CHD predominated, including ventricular septal defects (22/128), bicuspid aortic valve (9/128) and mild cardiomyopathy (3/128). Persistent truncus arteriosus was the most prevalent (n=8/13) conotruncal heart defect, but other anomalies also occurred in singleton cases. These findings support the need for cardiac evaluation in all individuals with distal 22q11.22-23 deletions, increased use of clinical genetic testing in syndromic individuals with these findings, and molecular studies in model systems. The results demostrate that reduced gene dosage of distal 22q11.21-23, particularly within the D-E region including *MAPK1* and *HIC2* convey risk for CHD.

## INTRODUCTION

The chromosome 22q11.21-22q11.23 region contains eight paralogous blocks of low copy repeats (LCR22s A-H) that are the main mediators of non-allelic homologous recombination events resulting in clinically relevant recurrent deletions across this region (Figure 1) ^1–4^. The deletion types have been named based upon the position of LCR22s with respect to the centromere, with LCR22-A being the most centromeric, and proximal along the q-arm as shown in Figure 1A (cen, centromere). Several genes in the 22q11.2 interval are illustrated for context (Figure 1A). While much is known about congenital heart disease (CHD) in individuals with deletions involving the proximal 22q11.21-22 region between LCR22-A and-D (A-D) that cause 22q11.2 deletion syndrome (22q11.2DS), where conotruncal heart defects (CTDs) predominate ^5–8^, there is relatively little known about the prevalence and spectrum of CHD in individuals with rarer recurrent distal 22q11.22-23 deletions involving the D-H region (Figure 1B-C).

**Figure 1.**
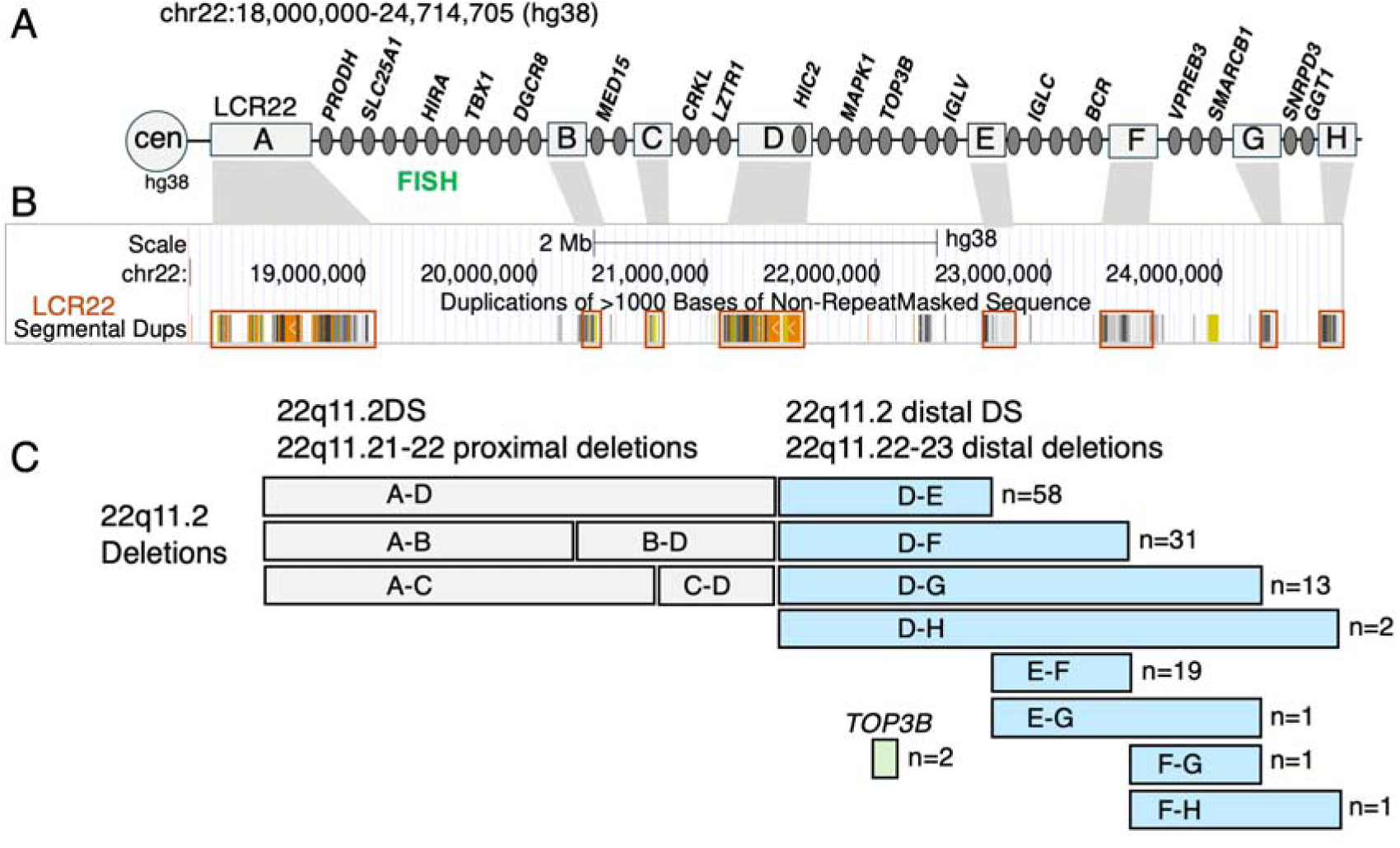
Physical map of the 22q11.21-22q11.23 region and sample sizes of distal deletions for the 128 total individuals studied. A) The interval on 22q11.2 is indicated in the hg38 assembly (the corresponding interval in hg19/GRCh37 is chr22:18,482,766-25,110,672), with the relative position of the centromere (cen) and eight blocks of low copy repeats shown (LCR22s lettered A-H, gray boxes), as well as the relative position and names of selected protein-coding genes for context purposes (dark gray circles). The genomic position of molecular fluorescence *in situ* hybridization (FISH) probes for identifying typical proximal deletions is indicated. B) Screenshot of the segmental duplication track (hg38) from the UCSC genome browser with correspondence to LCR22s A-H indicated above. C) Positions of recurrent deletions and respective syndrome names, across the 22q11.2 region, including the nine distal 22q11.22-23 deletions under study (eight flanked by LCR22s, blue boxes; TOP3B deletion, green box), together with their respective sample sizes, are indicated. DS = deletion syndrome.

The first individual with a distal 22q11.22-22q11.23 deletion was identified over 25 years ago, presenting with a complex conotruncal defect including interrupted aortic arch type B (IAAB) and persistent truncus arteriosus (PTA) with a D-F deletion ^9^. Since then, there have been several dozen individuals published in case reports or case series, and in previous reviews (Supplementary Table 1; ^10–14^). The clinical features of some of the individuals with distal deletions resemble those with a typical proximal deletion including conotruncal defects ^15,16^. It is therefore important to better delineate CHD types and frequencies in individuals with distal deletions.

Here we report the clinical cardiac findings for 128 unrelated individuals, including 62 newly reported, with distal 22q11.22-22q11.23 deletions. We found that CHD was present in 37.5% overall, a far greater prevalence than population-based expectations, but lower than that reported for typical proximal 22q11.2 deletions (A-D) ^17^, and with a pattern of decreasing prevalence of CHD for the most distal (and rarer) deletions in this LCR22-D to-H genomic interval. The CHD spectrum was generally milder than for typical proximal 22q11.2 deletions, and there were many additional individuals with congenital cardiac features that resolved spontaneously. A substantial minority of individuals had severe forms of CHD requiring surgery for survival. But of those with severe CHD, conotruncal defects were the most common and, PTA predominated. These results have important clinical implications for cardiac and genetic assessments and indicate a potential role of genes within the 22q11.22-23 distal deletion region, in particular the D-E region, which may contribute to risk for CHD.

## METHODS

### Consent, study design, participants and molecular genetic designation of 22q11.2 distal deletions

This was a retrospective study of unrelated individuals and is part of an IRB approved program at Albert Einstein College of Medicine (IRB #1999-201), Children’s Hospital of Philadelphia (IRB 07-5352), University Hospitals Leuven BE (IRB S52418), University of Toronto affiliated hospitals (IRB CAMH 114-2001, CAMH 154-2002, CAMH 150-2002, CAMH 151-2002 and UHN 98-E156), Sapienza (Protocol Ref. 5803) and Rome, IT (Prot. 468/12). These subjects were ascertained by their informed consent or assent. A waiver for formal approval under the Medical Research Involving Human Subjects Act (WMO) was obtained from the IRB of Amsterdam UMC, the Netherlands (#W20_098). All individuals in this study were de-identified for this study. Each person had a clinically relevant recurrent deletion in the 22q11.22-23 genomic region encompassed by LCR22 D-H, including recurrent deletions of *TOP3B*, which is a gene in the D-E region. *TOP3B* deletions are associated with neurodevelopmental disorders but not necessarily CHD ^18,19^. Newly reported individuals were clinically ascertained based on neurodevelopmental and/or syndromic findings (e.g. microcephaly, short stature, perauricular tags, dysmoprhic craniofacial features), with the genetic diagnosis made following clinical diagnostic testing. The majority of individuals were ascertained at the Children’s Hospital of Philadelphia, with the remainder contributed from Chromosome 22q11.2 Centers of Excellence in Belgium, Canada, Italy, and The Netherlands. All data was de-identified.

Previously published individuals with distal 22q11.22-23 deletions were procured through a detailed literature review using PubMed and the following search terms: ‘22q11 distal deletion’, with or without terms ‘heart’ and/or ‘congenital heart disease’, and from reports cited in prior publications, including those from previous reviews (Dykes et al., 2014b; Fagerberg et al., 2013). An addiitonal six individuals were included from reports published since the last review ^20,21^. We used similar inclusion and exclusion criteria for both newly described and previously published individuals in order to combine the cohorts for analyses in this study. However, individuals with unknown deletions sizes and/or microduplications included in previous studies were excluded from our analysis. Also, in contrast to previous reports, we included only unrelated individuals (i.e., excluding family members with the same distal deletion) because transmitting parents may present with milder disease. Additionally, we included those with sufficient cardiac assessment to determine CHD status.

The specific intervals harboring chromosome deletion breakpoints were determined using clinical genetic testing methods. These included MLPA (Multiplex Ligation-dependent Probe Amplification P250; MRC Holland), genome-wide/single nucleotide polymorphism-SNP microarray, whole exome sequencing, and (for previously published only) microsatellite dinucleotide genetic markers. All these methods made it possible to determine the approximate genomic extent of the deletions. We excluded individuals with unknown chromosome breakpoint intervals, or with a distal deletion and an additional genetic diagnosis including another pathogenic copy number variation, including those individuals whose deletion breakpoint regions were proximal to LCR22-D, such as LCR22 C-F, even if the distal deletion breakpoint was within the D-H region ^4^.

### CHD data obtained from individuals with distal 22q11.22-23 deletions

For the newly reported cohort, we compiled all genetic, demographic (that were available) and cardiac phenotypic data from the clinical reports. Data on congenital cardiac phenotypes were obtained from echocardiogram and clinical cardiology reports. For the previously published cases, we recorded comparable cardiac data from the prior phenotypic descriptions. All cardiac findings were reviewed by an experienced pediatric cardiologist.

We documented structural cardiac differences that were or would be observable prenatally, at birth or in early infancy. To stratify those with normal hearts and those with CHD, we considered all cardiac findings, including features that spontaneously resolved such as small muscular VSDs deemed to be hemodynamically insignificant (Supplementary Tables 1 and 2). Those with just one very mild cardiac feature found frequently in the general pediatric population were classified as a normal heart. Individuals with two or more of such defects (though each might be considered clinically insignificant on their own), and all individuals with clinically significant anomalies requiring intervention, were classified as having CHD. We assessed the severity of cardiac lesions based on current guidelines ^22–24^ and generated a consensus of severity rank of CHD (Supplementary Table 3).

### Statistical analysis

The main analyses calculated the prevalence of cardiac anomalies in individuals with deletions in the distal 22q11.22-23 genomic region, with 95% confidence intervals (95% CI) estimated using the normal distribution (Wald interval) formula; 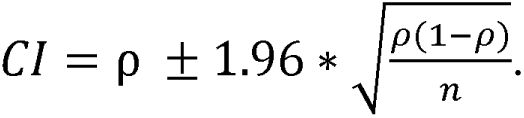 Confidence intervals were not calculated for sample sizes under four. We compared results between the newly reported and previously reported cohorts, to determine if there were differences, such as possible overrepresentation of cases with more severe phenotypes in older reports, that would preclude combining the cohorts. To assess differences in CHD patterns between distal and typical proximal 22q11.2 deletions (A-D), we compared findings for those individuals with distal deletions to those published previously with typical proximal 22q11.21-22 deletions ^17^. However, we did not have data on isolated bicuspid aortic valve (BAV), and therefore, this phenotype could not be compared. Categorical variables were compared using the Fisher’s exact test, with results reported as odds ratios with 95% CI and associated 2-tailed P-values. All statistical analyses were performed using SPSS software version 22 (SPSS, Chicago, IL) and R version 4.5.1. Differences were considered significant when the P-value was less than 0.05.

### Editorial Policies and Ethical Considerations

All individuals in this study were de-identified. There is not a key available that can be used to link de-identified data back to individuals from whom the data was collected.

## RESULTS

### Distal 22q11.22-23 deletion types with respect to position of LCR22s

The 22q11.2 region is associated with elevated likelihood of meiotic chromosomal misalignment mediated by LCR22s leading to deletions of various sizes and types, as is illustrated in Figure 1. The two main deletion categories involve proximal 22q11.21-22 deletions (predominantly A-D), and rarer, distal 22q11.22-23 deletions. Distal 22q11.22-23 deletions include copy number variants located between LCR22-D to-H (Figure 1A). These distal deletions are the focus of this report. Of clinical interest, distal deletions would not be detected using the standard clinical fluorescence in situ hybridization (FISH) testing with typical probes (N25 or TUPLE) commonly used to detect the typical proximal 22q11.21-22 deletion (Figure 1A).

In the cohort of 128 total individuals with distal 22q11.22-23 deletions, by far the most prevalent deletions had proximal endpoints in LCR22-D (104/128, 81.3%). The prevalence decreased based upon deletion extent: D-E (n=58, 45.3%), D-F (n=31, 24.2%), D-G (n=13, 10.2%), and D-H (n=2, 1.6%). The pattern was similar for deletions with proximal endpoints in LCR22-E: E-F (n=19,14.8%), E-G (n=1, 0.8%), and with the other three distal deletion types (F-G, F-H, *TOP3B*) involving only one or two individuals each (Figure 1C).

### Prevalence of CHD in individuals with different distal deletion types

A clinically relevant heart defect was present in 48 of the 128 individuals (37.5%, 95% CI: 29-46%) as shown in Table 1. There was no statistically significant difference in the overall prevalence of CHD between the new and previously reported cohorts (20/62, 32.3%, 95% CI: 20.6-43.9% vs 29/66; 43.9%, 95% CI: 31.9-55.9%; OR = 0.61, 95% CI: 0.29–1.28, p = 0.20; Supplementary Tables 1 and 2). We therefore combined the two cohorts (total n=128) for further analysis. Within the 80 individuals with normal hearts there were 20 (15.6% of the total sample) who had cardiac features present at birth that spontaneously resolved or required no clinical cardiology intervention or follow-up (Supplementary Table 2). Within individual distal 22q11.22-23 deletions, the prevalence of CHD was distributed as follows and illustrated in Figure 2: D-E (23/58, 39.7%, 95% CI: 27-53%), D-F (15/31, 48.4%, 95% CI: 31-66%), D-G (4/13, 30.8%, 95% CI: 6-56%), D-H (2/2, 100%), E-F (4/19, 21.1%, 95% CI: 3-39%), and none for the single individuals with E-G, F-G, and F-H deletions or the two with *TOP3B* deletions (Table 1). Considering just the 104 individuals with deletions with proximal endpoints in LCR22-D who represented the majority of the total sample (81.3%), there were 44 individuals with CHD (42.3%, 95% CI: 32.8-51.8%).

**Figure 2.**
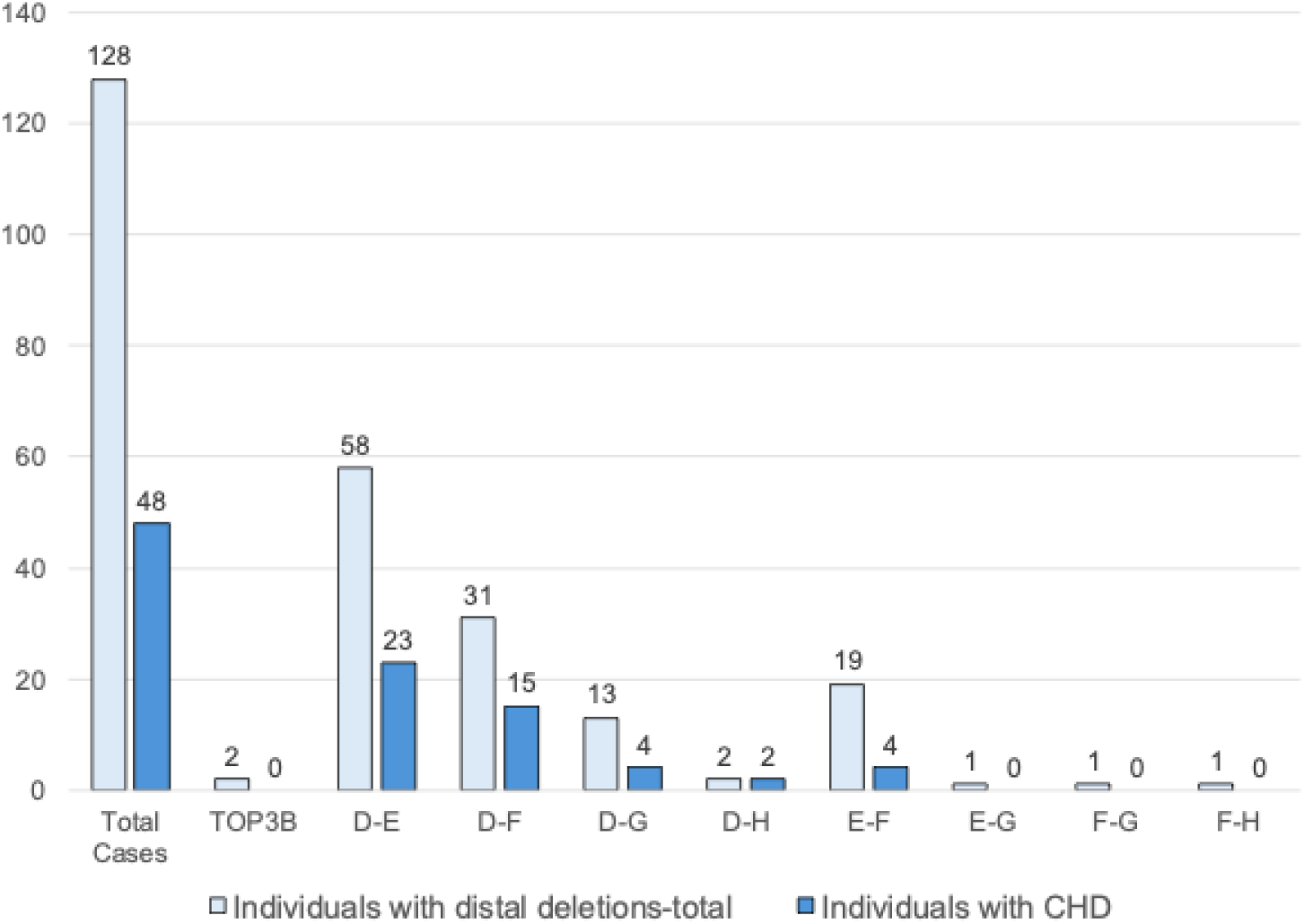
Bar graph of individuals with CHD among those with different distal deletion types. Number of individuals with distal deletions (gray bar) are indicated as are those with CHD (black bar) with different deletion types and numbers per group.

**Table 1.** Summary of cardiac lesions in individuals with distal 22q11.22-23 deletions sorted by deletion type. Abbreviations of cardiac phenotypes are provided below the table.

| Cardiac Lesion | Classification of CHD | Simplified Classification | Severity | TOP3B | D-E | D-F | D-G | D-H | E-F | E-G | F-G | F-H | Total Cases |
| --- | --- | --- | --- | --- | --- | --- | --- | --- | --- | --- | --- | --- | --- |
| VSD | VSD | VSD | Mild | - | 4 | 4 | 4 | 2 | - | - | - | - | 14 |
| ASD | ASD | ASD | Mild | - | - | 1 | - | - | 1 | - | - | - | 2 |
| VSD+ASD | VSD/ASD | VSD+ | Moderate | - | 2 | 1 | - | - | - | - | - | - | 3 |
| VSD+BAV | VSD/BAV | VSD+ | Moderate | - | 1 | - | - | - | - | - | - | - | 1 |
| VSD+Dextrocardia | VSD/Dextrocardia | VSD+ | Moderate | - | 1 | - | - | - | - | - | - | - | 1 |
| VSD+CoA | VSD/CoA | VSD+ | Moderate | - | 1 | 1 | - | - | - | - | - | - | 2 |
| VSD+MVI | VSD/MVI | VSD+ | Moderate | - | - | 1 | - | - | - | - | - | - | 1 |
| VSD+LVNC<br>(Cardiomyopathy)<br>+BAV | VSD/Cardiomyopathy/BAV | VSD+ | Moderate | - | - | 1 | - | - | - | - | - | - | 1 |
| BAV + LVNC<br>(Cardiomyopathy) | Cardiomyopathy/BAV | BAV+ | Moderate | - | - | 1 | - | - | - | - | - | - | 1 |
| BAV + Mild DCM<br>(Cardiomyopathy) | Cardiomyopathy/BAV | BAV+ | Mild | - | 1 | - | - | - | - | - | - | - | 1 |
| BAV | BAV | BAV | Mild | - | 3 | 1 | - | - | 1 | - | - | - | 5 |
| BAV+AS | AS/BAV | BAV+ | Moderate | - | - | 1 | - | - | - | - | - | - | 1 |
| Incomplete AVCD | AVCD | AVCD | Moderate | - | 1 | - | - | - | 1 | - | - | - | 2 |
| VSD+PS | Conotruncal | Conotruncal | Moderate | - | - | - | - | - | 1 | - | - | - | 1 |
| VSD+RAA | Conotruncal | Conotruncal | Moderate | - | 1 | - | - | - | - | - | - | - | 1 |
| PTA | Conotruncal | Conotruncal | Severe | - | 6 | 1 | - | - | - | - | - | - | 7 |
| PTA + IAAB | Conotruncal | Conotruncal | Severe | - | - | 1 | - | - | - | - | - | - | 1 |
| DORV | Conotruncal | Conotruncal | Severe | - | 1 | - | - | - | - | - | - | - | 1 |
| TOF | Conotruncal | Conotruncal | Severe | - | 1 | - | - | - | - | - | - | - | 1 |
| TGA+DILV | Conotruncal | Conotruncal | Severe | - | - | 1 | - | - | - | - | - | - | 1 |
| Patients w/ CHD |  |  |  | 0 | 23 | 15 | 4 | 2 | 4 | 0 | 0 | 0 | 48 |
| Patients w/<br>Deletion |  |  |  | 2 | 58 | 31 | 13 | 2 | 19 | 1 | 1 | 1 | 128 |
| Percent w/ CHD |  |  |  | 0 | 39.7 | 48.4 | 30.8 | 100 | 21.1 | 0 | 0 | 0 | 37.5 |

**Abbreviations:** Congenital heart disease (CHD), ventricular septal defect (VSD), ventricular septal defect and other lesions (VSD+), right sided aortic arch (RAA), atrial septal defect (ASD), bicuspid aortic valve (BAV), bicuspid aortic valve plus other defects (BAV+), left ventricular non-compaction (LVNC), dilated cardiomyopathy (DCM), persistent truncus arteriosus (PTA), interrupted aortic arch type B (IAAB), double outlet right ventricle (DORV), tetralogy of Fallot (TOF), transposition of the great arteries and double inlet left ventricle (TGA+DILV), pulmonic stenosis (PS), incomplete atrioventricular canal defect (AVCD), aortic stenosis (AS), mitral valve insufficiency (MVI), coarctation of the aorta (CoA).

We compared the frequency of CHD between individuals with distal deletions and those with typical proximal, 22q11.21-22, A-D deletions (Figure 1C). The overall prevalence of CHD in the individuals with distal 22q11.22-23 deletions was significantly lower than that reported for individuals with typical proximal 22q11.21-22 deletions (A-D; 650 of 1,182^17^, 55.0%, 95% CI: 51.3-59.0%; OR = 0.51, 95% CI: 0.35–0.73, p=3.7E-4).

### Distal deletion genotype correlations to CHD phenotype

The cardiac lesions, with their aggregate phenotypes arrayed by deletion type are shown in Table 1. We also classified them to five phenotype groups according to the main lesion type including ventricular septal defects (VSDs), atrial septal defects (ASDs) bicuspid aortic valve (BAV) and conotruncal heart defects as the main categories (Table 1). Figure 3 shows overall results for the five simplified CHD classifications, as well as normal hearts; Supplementary Tables 1 and 2 provide detailed data at the individual level.

**Figure 3.**
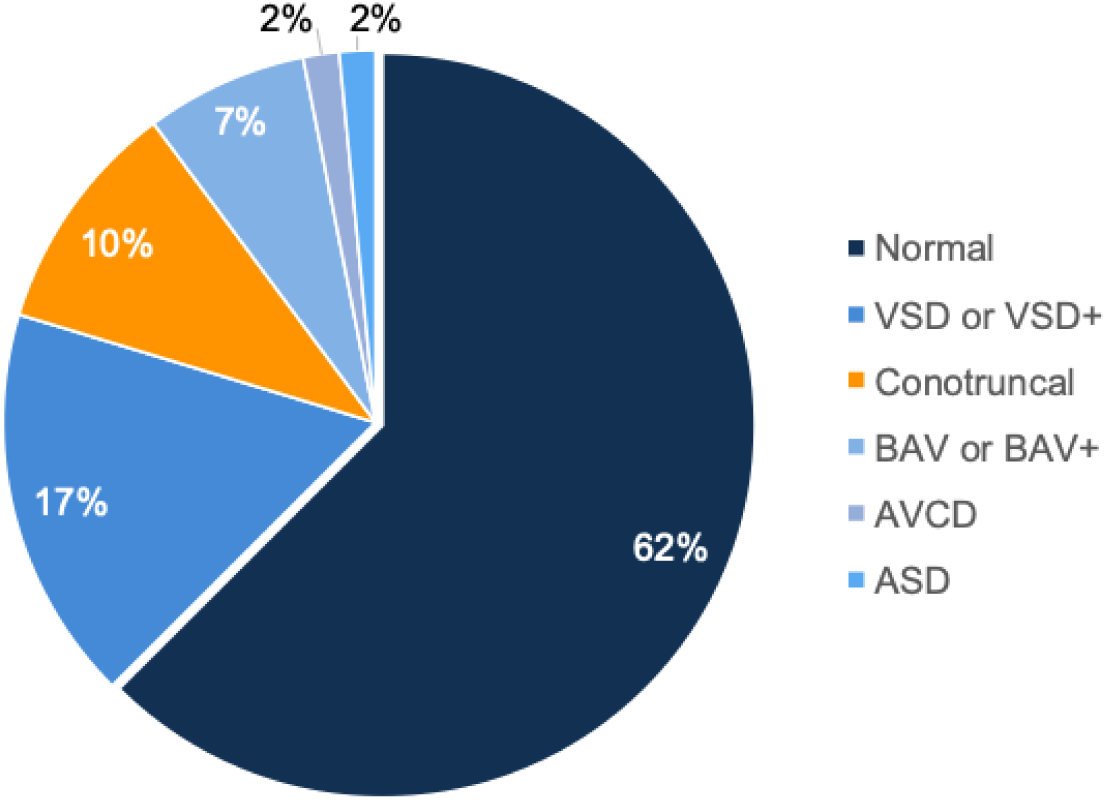
Relative prevalence of CHD types in the cohort of 128 individuals with distal 22q11.22-23 deletions. The five CHD classifications listed in Table 1 are displayed with respect to the percentage of the total individuals (n=128). More details for each individual in the study is provided in Supplementary Tables 1 (with literature citation for previously reported patients) and 2 (detailed cardiac phenotypes recorded for each individual of the 128 total).

The cardiac lesions were also parced into CHD severity groups from a consensus of current guidelines, including severe lesions requiring surgery for survival beyond early infancy ^22–24^. The majority (30 of 48, 62.5%) of individuals had CHD characterized as mild to moderate in severity (Table 1; Supplementary Table 3). The most frequent individual CHD type observed was an isolated VSD (n = 14, 11%, 95% CI: 6-16%). Septal defects including VSDs (mild muscular VSDs being more common), ASDs with/out other defects amounted to 52% (25 of 48 with CHD) of all CHD types in distal deletion individuals, excluding conotruncal defects (Table 1).

A total of 8.5% (11/128) had severe lesions and they were all conotruncal type defects. There were 13 individuals in total that had conotruncal defects comprising 10.2% (95% CI: 4.9-15.4%) of the total sample. Two of the 13 had less serious conotruncal defects (right sided aortic arch-RAA with a VSD; pulmonic stenosis-PS with a VSD; Table 1). The 13 total amounted to 27% (95% CI: 14.5-40.0%) of those with CHD. This prevalence is less than that reported for conotruncal defects in individuals with typical proximal 22q11.21-22 deletions (A-D; 461 of the total 1,182^17^, 39.0%; OR = 0.16, 95% CI: 0.09–0.29; RR = 0.24, 95% CI: 0.14–0.41; p = 7.1E-13) where they comprised 70.9% of the total of 650 individuals with CHD ^17^. Notably, in individuals with distal 22q11.22-23 deletions, PTA was most common conotruncal defect (n = 8 of 13 with conotruncal defects, 61.5%, 95% CI: 35-88%), with only one individual each with other complex conotruncal defects, such as TOF or IAAB (which occurred in conjunction with PTA; Table 1). While TOF, IAAB, and PTA characterize conotruncal defects in those with proximal deletions, TGA and DORV are rare. Therefore the relative proportion of types of conotruncal defects differ between the groups.

In addition to typical structural CHD types, cardiomyopathies, such as left ventricular non-compaction (LVNC), a previously suggested association in distal 22q11.22-23 deletions ^25^, and mild dilated cardiomyopathy, were reported in three individuals in our combined cohort (2.3% of the total, 6.3% of those with CHD; Table 1, Figure 3). Of note, the mild cardiomyopathy defects occurred in conjunction with BAV in all three individuals, and BAV overall was relatively prevalent, found in 8 of the 48 individuals with CHD (16.7%, 95% CI: 5.9-27.4%; 6.3% of the total sample, 95% CI: 2.1-10.4%; Table 1, Figure 3), greater than the general population frequency of BAV (0.5-2%) ^26^.

## DISCUSSION

Studying a combined cohort of 128 unrelated individuals with distal 22q11.22-23 deletions and data on CHD types, including 62 newly described individuals, has allowed us to add new insights into understanding genotype to CHD phenotype correlations for these rare deletions. First, it is clear that CHD is associated with distal 22q11.22-23 deletions, with the overall prevalence of 37.5% exceeding that of general population expectations by nearly 50-fold ^27^. Second, we determined that the spectrum of CHD is broad, with milder forms such as isolated septal defects, BAV and/or mild cardiomyopathy prevailing over more severe and complex conotruncal heart defects. Of note, the spectrum would have been even broader if we had included the many individuals with a single congenital cardiac feature that had resolved or had no clinical consequences, because these are common findings in the general normal population. Third, we delineated significant differences in CHD prevalence and spectrum or severity as compared with the more frequent proximal 22q11.2 deletions (A-B, A-C, A-D). Fourth, we established that the highest prevalence of CHD associated with distal 22q11.22-23 deletions is in the LCR22 D-E region. Collectively, these results have important clinical and research implications, for genetics and cardiology.

### Differing prevalence and spectrum of CHD in distal versus typical proximal 22q11.2 deletions

Appreciating differences between distal (D-H) and proximal (A-D) 22q11.2 deletions is essential for individual identification, genetic counseling, and understanding genomic pathways from deletion to phenotypic expression. The results of the current study demonstrate that the conotruncal defects that are a hallmark of CHD in the typical proximal 22q11.2 deletions (A-D) ^28–31^ are not only significantly less prevalent in individuals with distal 22q11.22-23 deletions but also have a different pattern of cardiac anomalies. Of note, the proportion of individuals with conotruncal defects in those with distal 22q11.22-23 deletions (10.2%) is actually comparable to that reported for the birth prevalence of these defects (0.12%, or <9%) in a Norwegian population-based study where 1.4% of children had CHD ^32^. Although this may suggest that conotruncal defects are not enriched within CHD types in distal 22q11.2 deletions, the details are paramount. Particularly striking is that PTA is the most frequent conotruncal lesion associated with distal 22q11.22-23 deletions, in contrast to the more common findings in proximal 22q11.2 deletions (A-D), such as TOF and IAAB, and with PTA representing fewer than 10% of those with CTDs ^17^. Equally important, in the general population, PTA is one of the rarest conotruncal defects, over an order of magnitude less prevalent than TOF, for example, suggesting that results for collective conotruncal defects may obscure differences in CHD patterns for distal deletions. These results therefore support clinical genetic testing for all individuals with PTA, assessing for deletions across the entire 22q11.2 deletion region, including the distal deletion region which would not be identified using the standard proximal FISH clinical diagnostic studies (N25 or TUPLE).

Other findings that appear to differ for distal 22q11.22-23 deletions include the elevated prevalence of BAV reported in one in every six individuals, and perhaps also LVNC/cardiomyopathy, anomalies not known to be associated with proximal deletions ^33,34^. Of interest, mouse models of abnormal NOTCH signaling indicate a possible connection between LVNC and BAV expression ^35^.

### Clinical implications

The results of this study clearly demonstrate that CHD is associated with distal 22q11.22-23 deletions, supporting the need for cardiac evaluations for all individuals, as early as possible, to help determine paths of clinical care. This is in the context of the lifetime complex care that is often required, given the other comorbidities that may be present or may develop, including prematurity, low birth weight, developmental delay, neurobehavioral and other medical conditions, including the enormously important potential for rhabdoid tumors in those with germline deletions including *SMARCB1,* located within the F-G region ^4,11,12,20,36,37^. For individuals with moderate to severe CHD, care would include standard cardiology follow-up, ideally by an expert team. More longitudinal data is needed to support routine cardiac follow-up for those with milder forms of CHD. However, we note that BAV is associated with late onset symptoms, and may present an elevated risk of aortic stenosis and/or aneurysm. Also, the possible association with LVNC, a condition which is often initially asymptomatic, suggests vigilance and a low threshold for cardiac imaging in individuals with distal 22q11.22-23 deletions. In all cases, long term management, incorporating knowledge of CHD associations, is required to optimize overall outcomes for individuals with the distal deletions described herein.

These findings also have important implications for cardiology. The CHD spectrum identified here suggests that previous recommendations for clinical genome-wide microarray or comparable testing for individuals with TOF or other conotruncal defects ^38^, should be applied to all those with PTA, at any age. This deserves emphasis as many healthcare providers assume clinical FISH testing with the typical proximal probe (Figure 1) would have detected any 22q11.2 deletion, including distal deletions. The association of BAV with distal 22q11.22-23 deletions in the current study likewise supports consideration of more widespread use of clinical genome-wide microarray or MLPA for milder forms of CHD in individuals with any other suggestive syndromic features such as short stature, microcephaly, preauricular tags, developmental delay, cognitive deficits, neuropsychiatric conditions, or other multi-system features.

### Genes implicated in cardiac development in the distal 22q11.22-23 deletion region

The most common distal 22q11.22-23 deletion identified in this combined cohort included the D-E region, and of those, 42% were associated with CHD. These results support the possibility that genes within the D-E region may play a critical role in early cardiac development sensitive to altered dosage. Foremost amongst these is *MAPK1*, a known candidate gene for CHD ^10,12,14,39,40^. *MAPK1* encodes a protein that mediates MAPK (Mitogen-activated protein kinase) signaling downstream of cell membrane receptor tyrosine kinase activation. Previous work has shown that mice with a conditional knockout of *MAPK1* in neural crest cells, which are a type of cardiac progenitor cell, had a phenotype including cleft palate and conotruncal defects ^41^. In addition, *HIC2*, located near the telomeric end of LCR22-D in a small region of unique, non-LCR22 sequences (Figure 1A and B), is deleted in most individuals with deletions overlapping the D-E region that involve a breakpoint in LCR22-D and in a small subset of individuals with a proximal 22q11.2 deletion ^42^. From studies in animal models, *HIC2* is implicated in the etiology of CHD as well ^42,43^. For other genes in the D-E region, including *TOP3B*, available data do not support evidence of a role in CHD.

In addition, the results suggest that genes in the E-F region may contribute to CHD risk. There may also exist position effects of these and other 22q11.22-23 deletions on neighboring genomic regions that could contribute to altered cardiac development.

### Strengths and limitations

The main strength of the current study is the addition of data on 62 new individuals with distal 22q11.22-23 deletions, nearly doubling the total number of individuals from those previously reported. The rarity of these deletions remains a major limitation. The population prevalence of these copy number variants is unknown, though estimates from biobanks suggest they are quite rare ^44^. It is possible that additional genomic testing using neonatal whole exome/genome sequencing may increase the likelihood of identifying individuals with a more severe types of CHD, or may decrease that likelihood, given the emphasis on clinical genetic testing for developmental delay. Also, we opted not to include family members with the same deletion, because they are likely to be less severely affected, especially in transmitting parents. The retrospective nature of the study precluded systematic detection of other contributing genetic factors for the sample studied. There were also limitations related to the lack of availability of echocardiography or cardiology reports for individuals reported in the literature. For example, some VSDs could not be distinguished as conoventricular requiring surgical repair versus muscular (mild), and these and other mild CHD such as LVNC and/or BAV, may have been missed in infants. Severe CHD would likely have been documented, however.

This work expands our understanding of congenital cardiac anomalies, including prevalence and spectrum of CHD associated with rare distal 22q11.22-23 deletions. Prevalence was lower and severity was less than in typical proximal 22q11.2 deletions (A-D). Milder forms of CHD, including septal defects, particularly VSDs, and BAV, predominated. PTA was by far the main conotruncal anomaly represented in the approximately 10% of individuals with these defects, with other severe CHD present in individual cases. Importantly, these results suggest that cardiac evaluations should be performed in all individuals identified with a distal 22q11.22-23 deletion, given the possibility of CHD that could have consequences across the lifespan. Other clinical implications include the need to consider referral for genetics consultation/genome-wide clinical genetic testing by cardiologists and perinatologists, for individuals with PTA especially amongst those with conotruncal defects, and more broadly for individuals with BAV, septal defects, and other milder forms of CHD in the presence of additional syndromic or developmental features. The findings also provide further support for the likelihood that genes in the distal deletion region, particularly within the D-E region including *MAPK1* and *HIC2*, may play a role in early cardiac development that is sensitive to altered gene dosage. Further research involving additional individuals with distal 22q11.22-23 deletions is needed to better define the full spectrum of CHD and long term outcome of these rare deletions, as well as to investigate modifier genes located elsewhere in the genome, acting in concert with effects of decreased dosage of genes in the distal deletion region, that may represent shared mechanisms with other pathogenic copy number of variations predisposing to CHD.

## Supporting information

Supplementary Table 1. Source publications and cardiac phenotype information for previously described individuals with 22q11.22-23 deletions. Each re

Supplementary Table 2. Cardiac phenotypes among 128 individuals with distal 22q11.22-23 deletions. Each individual in the study (column A) is listed

Supplementary Table 3. Cardiac lesion severity scores from three publications providing guidelines. The first author for each of three publications

## Data Availability

All data produced in this present work are contained in the manuscript.

## ACKNOWLEDGEMENTS

We owe an enormous debt of gratitude to all individuals and families who contributed to this investigation. This work was supported by NIH grants P01HD070454, R01HL157157, R01GM125757, R01GM125757-03, and U01MH101720, and in part by the Canadian Institutes of Health Research (CIHR) [MOP-313331 and MOP-111238] and from the Inaugural Dalglish Chair in 22q11.2 Deletion Syndrome at the University of Toronto and University Health Network to ASB.

## DECLARATION OF AI AND AI-ASSISTED TECHNOLOGIES IN THE WRITING PROCESS

AI was not used in the writing process.

## STATEMENT OF CONTRIBUTIONS

**Tanner J. Nelson:** Analyzed data and wrote the original draft at Albert Einstein College of Medicine, NY, USA.

**Daniel E. McGinn:** Human subject recruitment, phenotypic data acquisition, acquisition of biological specimens for confirmation of deletion size, quality control, informed consent, project administration, manuscript review and editing at Children’s Hospital of Philadelphia, PA, USA.

**Victoria Giunta:** Data entry, quality control, acquisition of biological specimens at Children’s Hospital of Philadelphia, PA, USA.

**Lydia Rockart:** Human subject recruitment, informed consent, acquisition of biological specimens at Children’s Hospital of Philadelphia, PA, USA.

**Audrey Green:** Human subject recruitment, informed consent, acquisition of biological specimens at Children’s Hospital of Philadelphia, PA, USA.

**Oanh Tran:** Performed DNA analysis to identify or confirm the 22q11.21 distal deletion at Children’s Hospital of Philadelphia, PA, USA.

**Daniella Miller:** Edited the manuscript and provided intellectual discussion at Albert Einstein College of Medicine, NY, USA.

**Elaine H. Zackai:** Performed medical and phenotypic analysis of patients, review and editing at Children’s Hospital of Philadelphia, PA, USA.

**T. Blaine Crowley:** Developed data dictionary, oversaw data management, IRB, project administration, and quality control at Children’s Hospital of Philadelphia, PA, USA.

**Beverly S. Emanuel:** Oversaw identification and confirmatory deletion size DNA analysis and edited the manuscript at Children’s Hospital of Philadelphia, PA, USA.

**Ann Swillen:** Recruited human subjects for this study from Center for Human Genetics, University Hospitals Leuven, Belgium and provided phenotypic data.

**Jeroen Breckpot:** Recruited human subjects for this study from Center for Human Genetics, University Hospitals Leuven, Belgium, and provided phenotypic data.

**M. Cristina Digilio:** Recruited human subjects for this study from Department of Medical Genetics, Bambino Gesù Hospital, Italy.

**Marta Unolt:** Recruited human subjects for this study from Department of Medical Genetics, Bambino Gesù Hospital and Department of Pediatrics, Gynecology, and Obstetrics, La Sapienza University of Rome, Italy.

**Carolina Putotto:** Recruited human subjects for this study from Department of Pediatrics, Gynecology, and Obstetrics, La Sapienza University of Rome, Italy.

**Federica Pulvirenti:** Recruited human subjects for this study from Department of Internal medicine, Endocrino-metabolic sciences and Infectious Diseases Referral Care Center for 22q11.2 Deletion Syndrome, Academic Hospital Policlinico Umberto I, Rome, Italy.

**Bruno Marino:** Recruited human subjects for this study from Department of Pediatrics, Gynecology, and Obstetrics, La Sapienza University of Rome, Italy.

**Zhengdong D. Zhang:** Performed data and statistical analyses for all comparisons.

**Elizabeth Goldmuntz:** Examined cardiac clinical data and assigned phenotype groups for this study at Children’s Hospital of Philadelphia, PA, USA.

**Erik Boot:** Data-inclusion and interpretation, writing-review and editing of the manuscript. **Anne S. Bassett:** Human subject recruitment and assessment, data-inclusion and interpretation, writing-review and editing of the manuscript.

**Bernice E. Morrow:** Edited the manuscript, generated figures and analyzed the data at Albert Einstein College of Medicine, NY, USA.

**Donna M. McDonald-McGinn:** Conceived the original study, oversaw human subject identification and recruitment, phenotypic data acquisition, acquisition of biologic specimens for confirmation of deletion size, quality control, informed consent, project administration including collaborative study, analyzed the data, and edited the manuscript at Children’s Hospital of Philadelphia, PA, USA.

## SUPPLEMENTARY TABLES

**Supplementary Table 1.** Source publications and cardiac phenotype information for previously described individuals with 22q11.22-23 deletions. Each reference citation is provided per row (column A). The institution and country of the last author is also provided support that these are different study subjects than the newly described ones (column B). We indicated the total individuals that fit our inclusion criteria (column C) and those reported with CHD (column D). We provided a breakdown of CHD versus deletion type (columns E-M) and indicated the cardiac severity as shown in the first row and basis for scoring is provided in detail in Supplementary Table 3). We provided a summary of each cardiac lesion per publication (column N) and the total number of individuals per deletion type (column O). More details on the cardiac lesions per individual is provided in Supplementary Table 2.

**Supplementary Table 2.** Cardiac phenotypes among 128 individuals with distal 22q11.22-23 deletions. Each individual in the study (column A) is listed by row. The cardiac history is provided (column B), as is the cardiac phenotype summaries that are color coded according to severity (column C; see Supplementary Table 3 for scoring criteria). Information about spontaneous closure of septal defects (column D) and/or that didn’t require further cardiology intervention (column E) is provided when available. We also included deletion type (column F), sex when available (column G), self reported race when available (column H) and ethnicity (column I. Heart anatomical terms are spelled out the first time they are mentioned (column A). NA means not available. Newly described individuals are listed first. Previouly published patients are listed below, as indicated. The first author of publications provided in Supplementary Table 1 are listed for individuals that were previously reported.

**Supplementary Table 3.** Cardiac lesion severity scores from three publications providing guidelines. The first author for each of three publications (with PubMed ID) is provided, with severity score listed. The references are also provided in the text of the manuscript. Missing data for particular lesion is indicated (not applicable, NA). The consensus of the severity scores that were used in this report is provided (column E) with the color coding (column F).

